# Evaluation of the Performance of the RECODe Equation with the Addition of Polygenic Risk Scores for Adverse Cardiovascular Outcomes in Individuals with Type II Diabetes

**DOI:** 10.1101/2023.05.03.23289457

**Authors:** Noah L. Tsao, Renae Judy, Michael G. Levin, Gabrielle Shakt, Regeneron Genetics Center, Penn Medicine BioBank, Benjamin F. Voight, Jinbo Chen, Scott M. Damrauer

## Abstract

**Aims/Hypothesis:** Individuals with T2D are at an increased risk of developing cardiovascular complications; early identification of individuals can lead to an alteration of the natural history of the disease. Current approaches to risk prediction tailored to individuals with T2D are exemplified by the RECODe algorithms which predict CVD outcomes among individuals with T2D. Recent efforts to improve CVD risk prediction among the general population have included the incorporation of polygenic risk scores (PRS). This paper aims to investigate the utility of the addition of a coronary artery disease (CAD), stroke and heart failure risk score to the current RECODe model for disease stratification.

**Methods:** We derived PRS using summary statistics for ischemic stroke (IS) from the coronary artery disease (CAD) and heart failure (HF) and tested prediction accuracy in the Penn Medicine Biobank (PMBB). A Cox proportional hazards model was used for time-to-event analyses within our cohort, and we compared model discrimination for the RECODe model with and without a PRS using AUC.

**Results:** The RECODe model alone demonstrated an AUC [95% CI] of 0.67 [0.62-0.72] for ASCVD; the addition of the three PRS to the model demonstrated an AUC [95% CI] of 0.66 [0.63-0.70]. A z-test to compare the AUCs of the two models did not demonstrate a detectable difference between the two models (p=0.97)

**Conclusions/Interpretation:** In the present study, we demonstrate that although PRS associate with CVD outcomes independent of traditional risk factors among individuals with T2D, the addition of PRS to contemporary clinical risk models does not specifically improve the predictive performance as compared to the baseline model.

**Research in Context:**

- Early identification of individuals with T2D who are at greatest risk of cardiovascular complications can lead to targeted intensive risk-factor modification with the aim of altering the natural history of the disease.
- Current approaches to risk prediction tailored to individuals with diabetes are exemplified by the RECODe algorithms which predict both individual and composite CVD outcomes among individuals with T2D.
- We sought to determine if the addition of a polygenic risk score to current clinical risk models improve predictive modeling of adverse cardiovascular events in individuals with type II diabetes.
- We demonstrate that although PRS associate with CVD outcomes independent of traditional risk factors among individuals with T2D, the addition of PRS to traditional, validated models does not specifically improve the predictive performance as compared to the base model.
- RECODe demonstrated modest discrimination potential at baseline (AUC = 0.66). As such, the lack of improved risk prediction may reflect the performance of the RECODe equation in our cohort as opposed to lack of PRS utility.
- Current performance of clinical risk models appears modest. Although PRS doesn’t meaningfully improve performance, there is still substantial opportunity to improve risk prediction.

## Introduction

Projected to impact up to 629 million people by 2045, Type-2 diabetes (T2D) is a leading cause of morbidity globally.^1^ Individuals with T2D are at increased risk of developing a wide range of cardiovascular complications.^2^ Early identification of individuals with T2D who are at greatest risk of cardiovascular complications can lead to targeted intensive risk-factor modification with the aim of altering the natural history of the disease. Current approaches to risk prediction tailored to individuals with diabetes are exemplified by the Risk Equations for Complications Of type II Diabetes (RECODe) algorithms which predict both individual and composite CVD outcomes among individuals with T2D. The RECODe were derived from the Action to Control Cardiovascular Risk in Diabetes (ACCORD) study and validated with the Diabetes Prevention Program Outcomes Study (DPPOS) and Look AHEAD (Action for Health in Diabetes).^3^

Recently, polygenic risk scores (PRS), which sum the genetic risk associated with common DNA variants across the genome have been shown to associate with complex diseases^4^ and have been posited to be useful is risk stratification.^4–12^ Despite this, there are conflicting data regarding their ability to improve clinical risk predication beyond current risk prediction algorithms.^4–14^

In this paper, we investigate the incremental benefit of PRS, beyond that of the RECODe equations, in improving the prediction of incident ASCVD events among individuals with T2D using data form a genomic medicine cohort recruited at a large academic medical center.

## Methods

Study approval was obtained from local institutional review boards and all patients enrolled in the PMBB provided written informed consent.

### Data Sources

The Penn Medicine Biobank (PMBB) is an academic healthcare system based genomic and precision medicine cohort (> 60,000 individuals at time of data extraction, July 2020) that links participant blood and tissue samples with associated health information. Procedures for recruitment, consent, data collection and genotyping are detailed elsewhere.^15^

Three publicly available genome wide association studies (GWAS) were used for PRS generation. The heart failure (HF) polygenic risk score was calculated using a GWAS from the Levin et. al comprised of over 115,000 HF individuals with all cause HF.^16^ The coronary artery disease (CAD) polygenic risk score was calculated using a meta-analysis from the Million Veterans Program, CARDIoGRAMplusC4D, the UK BioBank (UKBB) and Biobank Japan (BBJ)^17^ comprised of over 243,000 individuals with CAD. GIGASTROKE is a large-scale international collaboration^18^ that analyzed data comprised of over 110,00 individuals with ischemic stroke.

### Population

We created a prospective cohort study of incident ASCVD events following T2D diagnosis among PMBB participants with T2D. Diagnosis of T2D and ASCVD were defined based on a combination of electronic health record (EHR) diagnosis/procedure codes, medication prescriptions and laboratory measurements (**Supplemental Table 1**). Cohort participants were required to have at least 1 year of EHR data following their diagnosis of T2D and genome-wide genotype data. The date of initial T2D diagnosis served as the baseline date of entry to the cohort.

Individuals were excluded from the cohort if they had baseline history of ASCVD, defined as the presence of a diagnostic code for any of the following: (1) stroke or cerebrovascular disease, including hemorrhagic stroke, ischeic stroke, and transient ischemic attack; (2) coronary heart disease or coronary artery disease, including myocardial infarction (MI), angina, and coronary insufficiency (**Supplementary Table 2)**. The RECODe equation was optimized based on self-reported race, with a particular focus on White and Black individuals. In contrast, polygenic risk scores typically focus on subpopulations which share genetic ancestry.

Here, we focused on subpopulations with genetic similarity to the 1000 Genomes European- and African-ancestry superpopulations, based on genetic principal components.

### Outcomes

The primary outcome was incident atherosclerotic cardiovascular disease (ASCVD), defined as fatal or nonfatal ischemic stroke, fatal or non-fatal MI, or death from cardiovascular cause. Secondary outcomes included the individual outcomes of MI, stroke, congestive heart failure (CHF) or death from any cardiovascular cause. Vital status and date and cause of death, if applicable, were derived from a combination of EHR and National Death Index data. ICD codes for all primary and secondary outcomes can be found in the Data Supplement (**Supplemental Table 2)**.

### Risk Factors for ASCVD

Based on the RECODe model, we considered the following traditional risk factors as in the calculation of 10-year composite ASCVD risk: age, sex, ethnicity, current tobacco smoking, systolic blood pressure, history of cardiovascular disease, statin use, use of anticoagulants, use of anti-hypertensives, Hb-A1C, total cholesterol, serum creatinine and the urine albumin:creatinine ratio. Age was defined as age at initial T2D diagnosis, sex was defined as biological sex and derived from the EHR, race/ethnicity was defined as genetic similarity to EUR or AFR superpopulations.^19,20^ We used genetic similarity to the EUR and AFR superpopulations as opposed to RECODe’s EHR defined race/ethnicity as EHR data has been shown to inaccurately capture race/ethnicity and to appropriately model PRS.^20–22^ Statin use, Anti-coagulant use, and Anti-Hypertensive use were defined by having any one of the prescriptions listed in **Supplemental Table 3**. Consistent with RECODe, smoking history was defined as current or non-current (a composite of never and former smoking). Laboratory values were taken within 30 days of each patient’s T2D diagnosis and all values outside of 3 median absolute deviations from the median were treated as missing.

### Polygenic Risk Score Creation

PRS for CAD, ischemic stroke, and heart failure were developed based on GWAS summary statistics from Tcheandijeu et. al, the GIGASTROKE consortia and Levin et. al respectively.^16–18^ To reduce the total SNP set to a size amenable for PRS analysis, we extracted SNPs present in the HapMap 3+ reference panel (n =1,444,196 SNPs in HapMap panel; retained 1,099,988 SNPs in the CAD GWAS, 982,552 SNPs in the stroke GWAS and 1,122,273 SNPs in the HF GWAS). LDPred (v 2.0) (https://github.com/privefl/paper-ldpred2) was used to generate the posterior mean effect of each SNP based on LD information from the HAPMAP 3+ reference panel. Participant level PRS values were calculated using PLINK v2.0 (https://www.cog-genomics.org/plink). The individual PRS were scaled separately among AFR and EUR populations prior to all analysis.

### Statistical Analysis

The analytic cohort was split into training (80%) and validation sets (20%) at random. This was done to avoid overfitting given the 20 models tested, (3 PRS, a model containing no PRS, a model containing all three PRS and 4 outcomes). The training data was used to generate a RECODe model with the addition of PRS and to test the association of the various PRS with their outcome of interest. In this newly generated model, the coefficients for each of the risk factors included in the original RECODE equation retained their original values, however, the training data was used to calculate a coefficient for various PRS. The validation set was used to compare the discrimination and calibration of the RECODe model with and without PRS.

In the training dataset, we fit Cox proportional hazard models containing the RECODe risk factors and each of the PRS, both individually and together, to model both primary and secondary outcomes for a total of 20 different models (**Supplemental Table 9-13**). We fixed the weights on the RECODe variables at those specified by the original paper and used the model to determine the appropriate weights for the PRS terms. Individuals without primary or secondary outcomes were censored to the date of their last EHR encounter or non-CVD related death. The proportional hazard assumptions were tested based on Grambsch and Therneau’s proportional hazards tests and diagnostics.^23^ Missing data was imputed using the multiple chain imputation method as implemented by the MICE package in R.^24^

In the validation dataset, discrimination was assessed based on the receiver operator curve, and through calculating the area under the curve (AUC) as defined by Pepe et. al. AUCs, with bootstrapped 95% confidence intervals calculated using the survAccuracyMeasures package in R.^25,26^ To calculate these measures, high risk was defined as 20% or greater risk as per the ACC/AHA guidelines.^27^ AUC between models were compared using a z-test. Calibration was evaluated by comparing model predicted ASCVD to observed outcomes based on unadjusted Kaplan Meir statistics to account for unequal follow-up time. We also compared models based on the proportion of cases followed [PCF(q)], which represents the proportion of individuals who will develop disease who are included in the proportion q of individuals at highest risk, and the proportion needed to follow-up [PNF(p)], which represents the proportion of the population at highest risk that needs to be followed to identify the proportion p of individuals who will develop disease.

All analyses we performed in R version 3.6.2. Statistical significance was set at two sided α = 0.05.

## Results

### Cohort Demographics

Of the 63,104 individuals enrolled in the Penn Medicine Biobank at the time of data extraction, there were 10,528 with T2D. Of these, 3,588 were excluded because they had CVD events prior to their T2D diagnosis and 2,597 because they did not have genotype data available. This left 4,344 (6.87%) participants who met the cohort inclusion criteria **(Table 1)**.

**Table 1.** Cohort Demographics

|  | Validation | Training | P-value | Overall |
| --- | --- | --- | --- | --- |
|  | (N=868) | (N=3476) |  | (N=4344) |
| Age |  |  |  |  |
| Mean (SD) | 63.8 (13.5) | 63.9 (13.2) | 0.884 | 63.9 (13.2) |
| Median [IQR] | 65.4 [55.8, 72.9] | 65.0 [56.6, 72.8] |  | 65.1 [56.4, 72.8] |
| Gender |  |  |  |  |
| Female | 434 (50.0%) | 1731 (49.8%) | 0.946 | 2165 (49.8%) |
| Male | 434 (50.0%) | 1745 (50.2%) |  | 2179 (50.2%) |
| Genetic Similarity |  |  |  |  |
| African | 430 (49.5%) | 1756 (50.5%) | 0.633 | 2186 (50.3%) |
| European | 438 (50.5%) | 1720 (49.5%) |  | 2158 (49.7%) |
| BMI (kg/m2) |  |  |  |  |
| Mean (SD) | 32.9 (7.09) | 32.9 (7.33) | 0.953 | 32.9 (7.28) |
| Median [IQR] | 32.0 [28.0, 37.0] | 32.0 [28.0, 37.0] |  | 32.0 [28.0, 37.0] |
| Smoking Status (Ever/Never) |  |  |  |  |
| Yes | 864 (99.5%) | 3464 (99.7%) | 0.849 | 4328 (99.6%) |
| No | 4 (0.5%) | 12 (0.3%) |  | 16 (0.4%) |
| Anti-Hypertensive Medication |  |  |  |  |
| Yes | 793 (91.4%) | 3143 (90.4%) | 0.433 | 3936 (90.6%) |
| No | 75 (8.6%) | 333 (9.6%) |  | 408 (9.4%) |
| Statin |  |  |  |  |
| Yes | 679 (78.2%) | 2773 (79.8%) | 0.335 | 3452 (79.5%) |
| No | 189 (21.8%) | 703 (20.2%) |  | 892 (20.5%) |
| Systolic Blood Pressure (mmHg) |  |  |  |  |
| Mean (SD) | 131 (17.1) | 131 (17.2) | 0.658 | 131 (17.2) |
| Median [IQR] | 130 [120, 142] | 130 [120, 142] |  | 130 [120, 142] |
| Diastolic Blood Pressure (mmHg) |  |  |  |  |
| Mean (SD) | 75.7 (11.1) | 75.3 (11.1) | 0.452 | 75.4 (11.1) |
| Median [IQR] | 76.0 [69.0, 83.0] | 75.0 [68.8, 82.0] |  | 75.0 [69.0, 82.0] |
| Total Cholesterol (mg/dL) |  |  |  |  |
| Mean (SD) | 164 (40.5) | 163 (41.1) | 0.391 | 163 (41.0) |
| Median [IQR] | 161 [136, 190] | 160 [133, 190] |  | 161 [134, 190] |
| HDL Cholesterol (mg/dL) |  |  |  |  |
| Mean (SD) | 47.4 (13.9) | 46.7 (13.8) | 0.208 | 46.8 (13.8) |
| Median [IQR] | 46.0 [37.0, 56.0] | 45.0 [37.0, 55.0] |  | 45.0 [37.0, 55.0] |
| Max A1C (mg/dL) |  |  |  |  |
| **Mean (SD)** | 7.13 (1.27) | 7.01 (1.28) | 0.0084 | 7.03 (1.28) |
| **Median [IQR]** | 6.90 [6.20, 7.80] | 6.70 [6.10, 7.70] |  | 6.75 [6.10, 7.70] |
| **Serum Creatinine** |  |  |  |  |
| **Mean (SD)** | 1.00 (0.314) | 1.03 (0.325) | 0.0382 | 1.02 (0.323) |
| **Median [IQR]** | 0.950 [0.780, 1.14] | 0.970 [0.790, 1.21] |  | 0.970 [0.790, 1.20] |
| **Urine albumin:creatinine ratio, mg/g** |  |  |  |  |
| **Mean (SD)** | 1.89 (1.89) | 1.84 (1.94) | 0.559 | 1.85 (1.93) |
| **Median [IQR]** | 1.15 [0.606, 2.47] | 1.08 [0.556, 2.48] |  | 1.09 [0.566, 2.48] |
| **History of CVD** |  |  |  |  |
| **Yes** | 767 (88.4%) | 3116 (89.6%) | 0.302 | 3883 (89.4%) |
| **No** | 101 (11.6%) | 360 (10.4%) |  | 461 (10.6%) |
| **Anti Coagulant Use** |  |  |  |  |
| **Yes** | 491 (56.6%) | 1971 (56.7%) | 0.973 | 2462 (56.7%) |
| **No** | 377 (43.4%) | 1505 (43.3%) |  | 1882 (43.3%) |

Among the analytic cohort of 4,344 participants, the median [IQR] index age was 65.1 [56.4-72.8] years, 2,179 (50.2%) were men and 2,186 (50.3%) were genetically similar to AFR reference populations. Table 1 shows the baseline characteristics of this cohort. Overall, 4,328 patients (99.6%) were smoking at baseline, 3,452 (79.5%) had a prescription for statin therapy, and 3,936 (90.6%) had a prescription for anti-hypertensive medication **(Table 1)**.

Over a median follow up time of 7.0 years (IQR 2.9-11.1) and a total of 33,742.9 person-years of follow-up, 1,300 (30.0%) participants experienced the composite ACVD outcome and 1,941 (44.1%) experienced secondary outcomes including: 970 (22.3%) that had incident CAD, 575 (13.2 %), that had incident CVD, 873 (20.1%) that had incident HF, and 55 (1.3%) that died from a CVD event (**Figure 1, Supplemental Figure 1-4, Table 2**).

**Figure 1.**
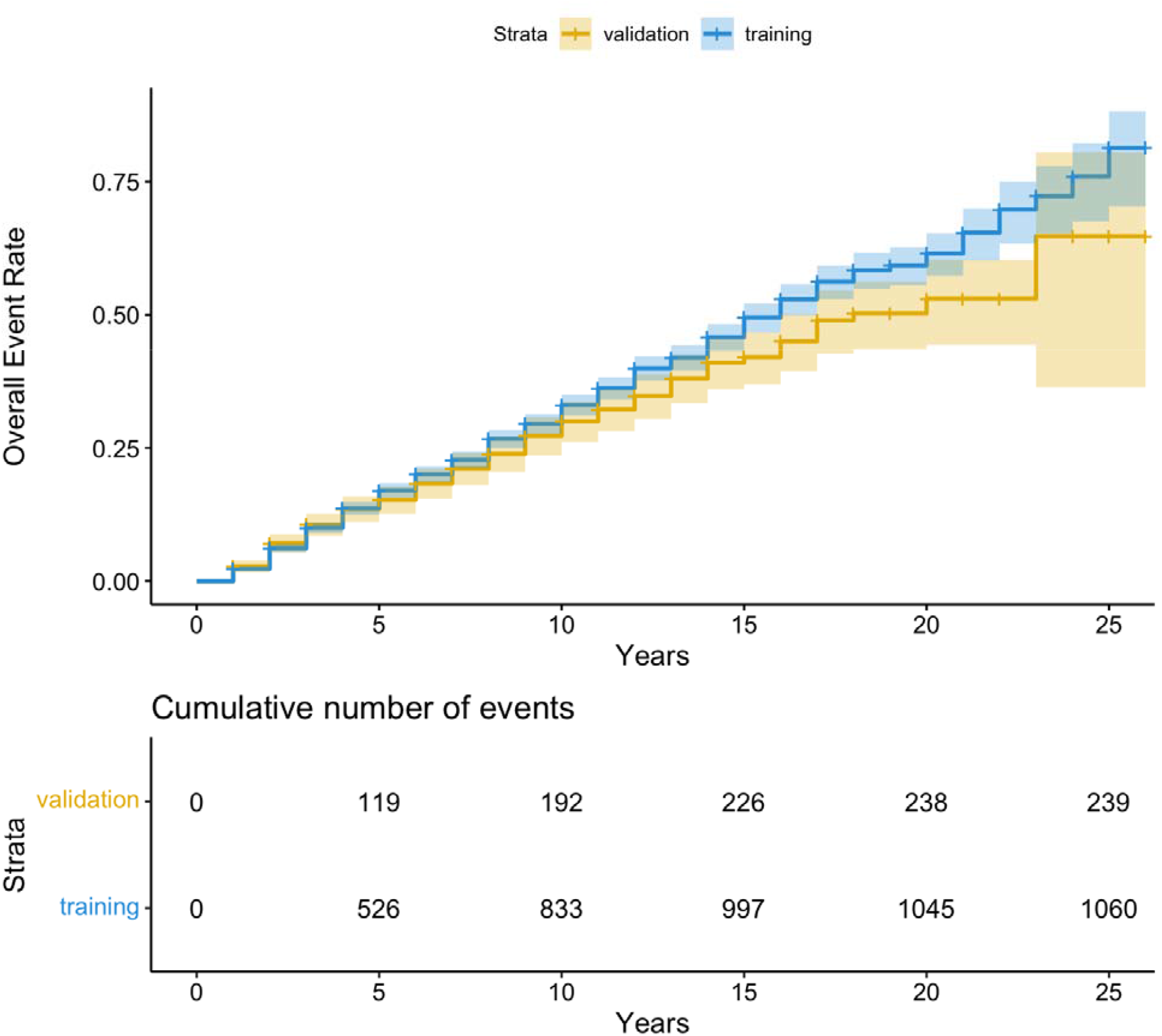
Unadjusted ASCVD Event Rate Curve of event rate data to ASCVD event (in months) after the diagnosis of T2D. The number of events at 5, 10, 15, 20 and 25 years are shown in a table immediately below the survival curves.

**Table 2.** Cohort Outcome Statistics

|  | Validation | Training | P-value | Overall |
| --- | --- | --- | --- | --- |
|  | (N=868) | (N=3476) |  | (N=4344) |
| MI |  |  |  |  |
| Yes | 101 (11.6%) | 400 (11.5%) | 0.963 | 501 (11.5%) |
| No | 767 (88.4%) | 3076 (88.5%) |  | 3843 (88.5%) |
| HF |  |  |  |  |
| Yes | 174 (20.0%) | 699 (20.1%) | 0.875 | 873 (20.1%) |
| No | 604 (69.6%) | 2379 (68.4%) |  | 2983 (68.7%) |
| Missing | 90 (10.4%) | 398 (11.5%) |  | 488 (11.2%) |
| CAD |  |  |  |  |
| Yes | 192 (22.1%) | 778 (22.4%) | 0.904 | 970 (22.3%) |
| No | 676 (77.9%) | 2698 (77.6%) |  | 3374 (77.7%) |
| Stroke |  |  |  |  |
| Yes | 99 (11.4%) | 476 (13.7%) | 0.0848 | 575 (13.2%) |
| No | 769 (88.6%) | 3000 (86.3%) |  | 3769 (86.8%) |
| Composite CVD |  |  |  |  |
| Yes | 239 (27.5%) | 1060 (30.5%) | 0.0964 | 1299 (29.9%) |
| No | 629 (72.5%) | 2416 (69.5%) |  | 3045 (70.1%) |
| Death by CVD Event |  |  |  |  |
| Yes | 5 (0.6%) | 50 (1.4%) | 0.0624 | 55 (1.3%) |
| No | 863 (99.4%) | 3426 (98.6%) |  | 4289 (98.7%) |

### Polygenic Risk Scores

Polygenic risk score optimization in the overall PMBB cohort was performed by LDpred2, and excluded individuals in the validation cohort. We tested the association of the HF, CAD and stroke PRS with their prevalent outcomes in both the overall PMBB cohort and in the T2D training cohort used in the following analyses. The HF PRS demonstrated a robust association with prevalent heart failure in both the overall cohort (OR 1.14 [95% CI 1.1-1.19]) as well as the T2D training cohort (OR 1.17 [95% CI 1.05-1.31]). Although both the CAD and stroke PRS demonstrated robust associations with their prevalent outcomes in the overall cohort (OR 1.41 [95% CI 1.37-1.45], OR 1.12 [95% CI 1.09-1.16] respectively), they failed to demonstrate an association in the T2D training cohort (OR 1.08 [95% CI 0.99-1.17], OR 1.08 [95% CI 0.98-1.21]).

We first sought to understand the association of the CAD, stroke and HF polygenic risk scores when added individually to RECODe risk models for either their individual incident outcomes, CVD-Death, or composite ASCVD when tested during model creation in the training dataset. Few PRS demonstrated statistically significant associations with any of the outcomes of interest. **(Supplemental Tables 4-8)**. There was a statistically significant association between the stroke PRS and ASCVD (HR 1.08 [95%CI 1.01 -1.14]) and the stroke PRS with incident HF (HR 1.09 [95%CI 1.01-1.17]), when added to the RECODe model for each outcome respectively **(Supplemental Tables 4, 8)**. Following this, we created models for each outcome of interest (ASCVD, CVD-Death, MI, stroke and HF) that included all three PRS tested. The only associations that demonstrated statistically significant results were the stroke PRS with composite ASCVD (OR 1.07 [0.95% CI 1.01-1.13] and the stroke PRS with incident HF (OR 1.09 [0.95% CI 1.01-1.17]. (**Supplemental Table 4**,**8)**.

### Utility of adding PRS to RECODe for risk prediction ASCVD

To assess the utility of the addition of a CAD, stroke and HF PRS to the RECODe equation for ASCVD, we first tested the discrimination of the models using time dependent AUC based on receiver operator curves **(Figure 2)**. The RECODe model alone demonstrated an AUC [95% CI] of 0.66 [0.62-0.71] and the addition of the three PRS to the model demonstrated an AUC [95% CI] of 0.66 [0.62-0.70]. A z-test to compare the AUCs of the two models demonstrated no difference in model discrimination with a p-value of 0.97 **(Supplemental Table 9)**. Calibration was visualized by calculating the projected cumulative event rate based on unadjusted Kaplan Meyer model in each decile of risk (**Figure 3**). The raw RECODe equation demonstrated a slightly more consistent decrease in survival probability over consecutive deciles of risk, but not in a statistically significant manner. Finally, we compared the PCF(q) (proportion of individuals that develop disease who are included in the proportion q of individuals at highest risk) and PNF(p) (proportion of the population at highest risk that needs to be followed to identify the proportion p of individuals who will develop disease) curves for each model. The PCF(q) and PNF(p) curves models including and not including the PRS are comparable **(Figure 4-5)**.

**Figure 2.**
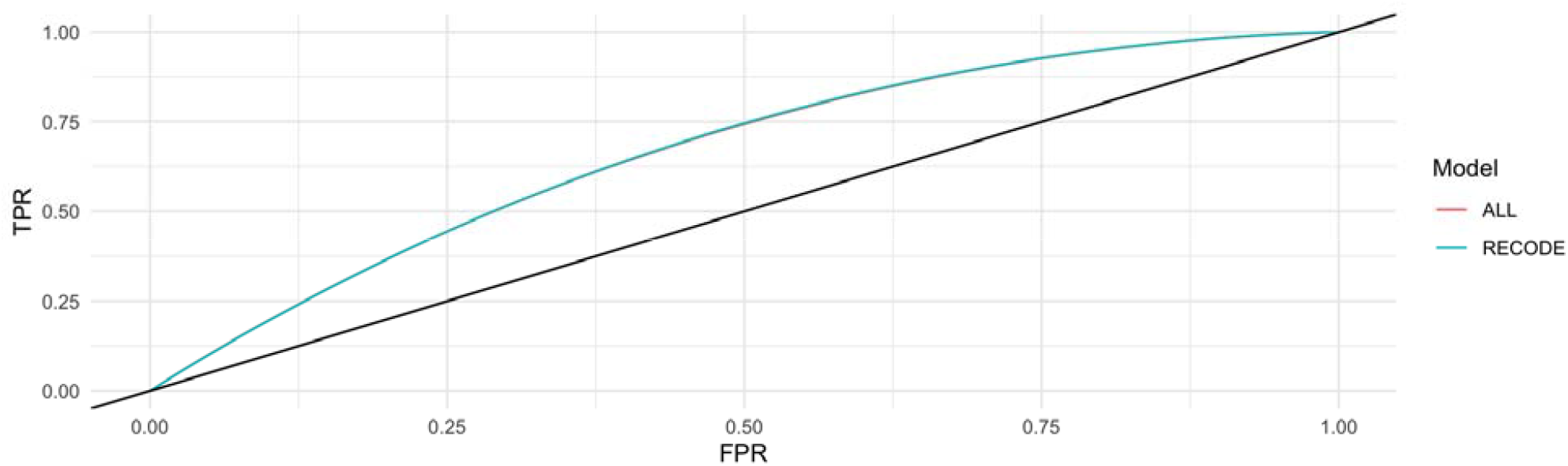
AUC of the RECODe Model for ASCVD with and without the addition of the PRS. The blue curve illustrates the AUC curve of the cox model of the RECODe model without the addition of the PRS. The red curve illustrates the AUC of the cox model of the RECODe model with the addition of a CAD, stroke and HF PRS.

**Figure 3.**
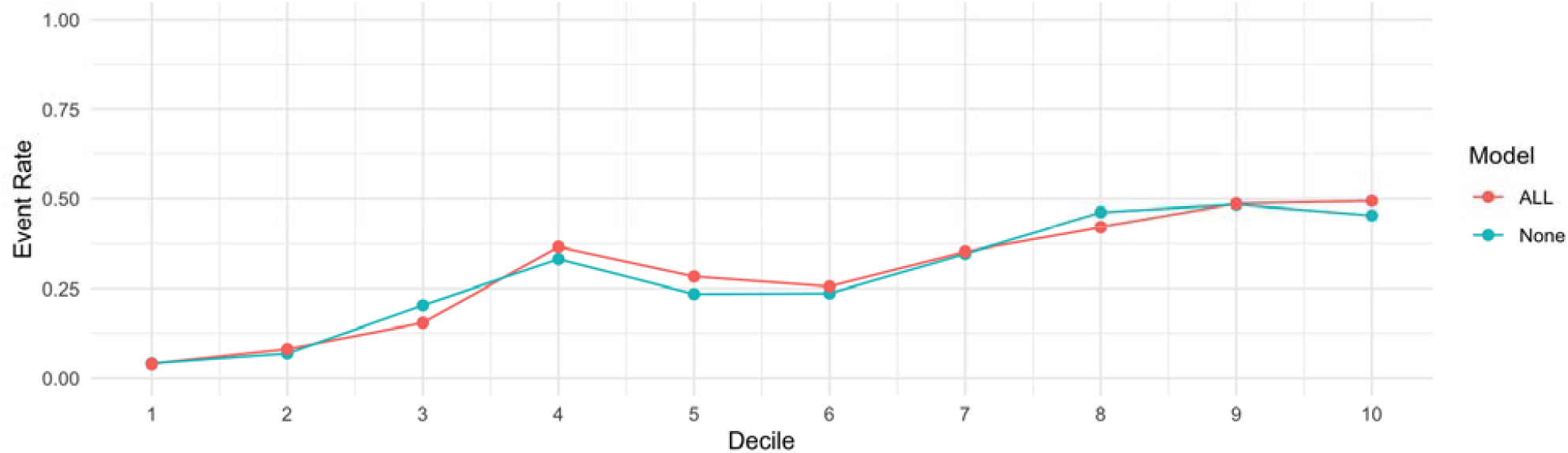
Event Rate Across Deciles of Risk between the RECODe Model for ASCVD and RECODe model with the addition of the PRS. The blue curve illustrates the cumulative event rate at each decile of risk based on an adjusted Kaplan Meyer model based on the RECODe model without the addition of the PRS. The red curve illustrates the same event rate curve of the RECODe model with the addition of a CAD, stroke and HF PRS.

**Figure 4.**
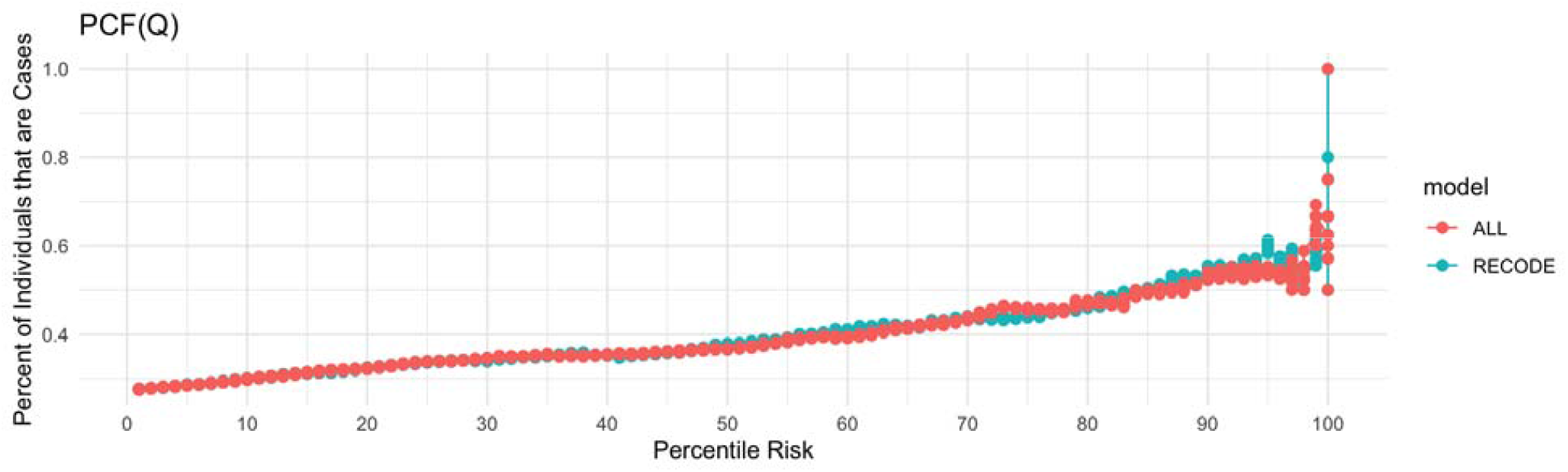
PCF(q) plot for the RECODe Model for ASCVD with the addition of the PRS. PCF(q) is the proportion of individuals who will develop disease who are included in the proportion q of individuals in the population at highest risk. The blue curve illustrate the PCF(Q) for the unadjusted RECODe model whereas the red curve illustrates the PCF(Q) for the RECODe model with the addition of a CAD, stroke and HF PRS.

### Secondary Outcomes

We also tested model performance for the addition of PRS to the RECODe model in four secondary outcomes: cardiovascular mortality, myocardial infarction (MI), ischemic stroke (IS) and Heart Failure (HF). Each of these secondary outcomes was modeled by the same risk factors described previously as per the RECODe equation. Model performance was evaluated using the methods described above for discrimination and calibration. Across all secondary outcomes, the addition of the PRS did not improve model discrimination or calibration by AUC, survival probability visualization, PCF(q), and PNF(p) analysis **(Supplemental Figures 5-8 and Supplemental Tables 10-13)**

**Figure 5.**
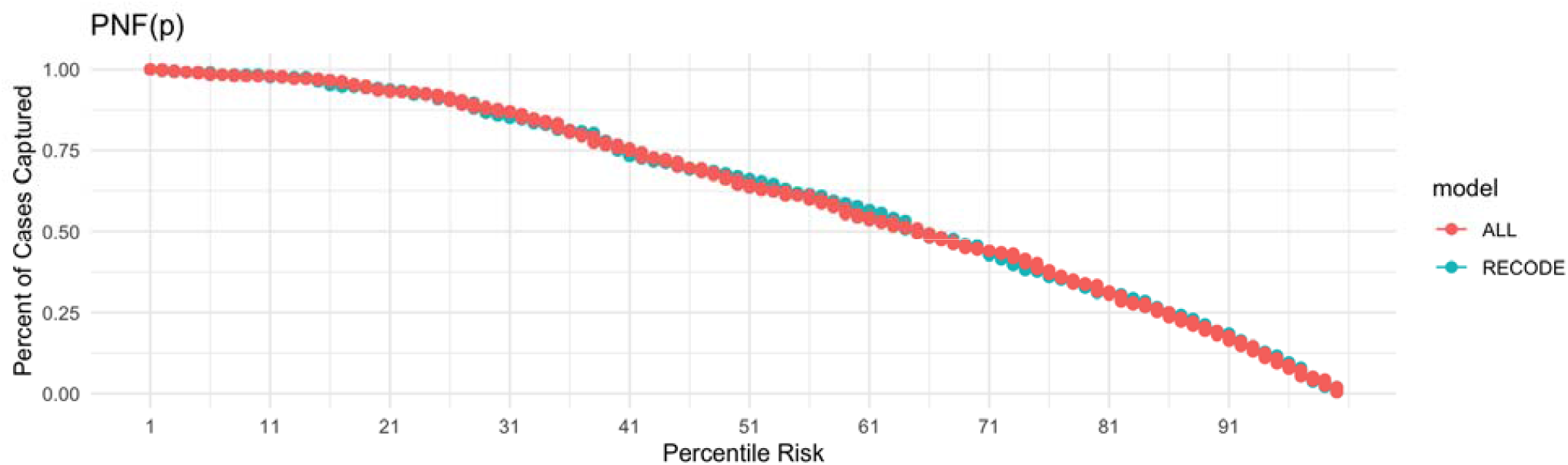
PNF(p) plot for the RECODe Model with the addition of the CAD, stroke and HF PRS. PNF(p) represents the percentage of individuals that we will capture if we follow individuals that are at the xth percentile of risk or higher. The blue curve illustrate the PNF(p) for the unadjusted RECODe model whereas the red curve illustrates the PNF(p) for the RECODe model with the addition of a CAD, stroke and HF PRS.

## Discussion

In the present study, we demonstrate that although PRS associate with CVD outcomes independent of traditional risk factors among individuals with T2D, the addition of PRS to clinical risk models does not specifically improve the predictive performance as compared to the base model. This was true across all cardiovascular disease outcomes tested and across various discrimination and validation techniques, suggesting that the addition of PRS would also not improve risk stratification in our cohort. Our findings among individuals with T2D are consistent with those reported for the population at large and corroborate a study by Mosely et al. in which they showed no significant improvement in reclassification of cardiovascular risk with models including both the PCE and a CAD PRS versus PCE alone despite the association of their CAD PRS with incident disease in the ARIC and MESA studies.^28^

These results add to the growing body of literature suggesting that the relative additional value of PRS for risk prediction among individuals in whom the clinical risk factors are both manifest and measurable is likely modest to negligible. Interestingly, in our study the RECODe equation showed discrimination potential at baseline (AUC = 0.66). As such, the lack of improved risk prediction may reflect the performance of the RECODe equation in our cohort as opposed to lack of PRS utility.^29^ In other words, the performance of the RECODe equation alone may have hit the ceiling of predictive performance in our cohort. This could be due to overrepresentation of older individuals and individuals with cardiovascular disease in our cohort as opposed to the general population.

It has been proposed that PRS may have a role in predicting complex diseases early in a person’s life course before traditional clinical risk factors may have had a chance to manifest, but may have less clinical utility in individuals that have a history in the healthcare system.^30^ Given the median age of onset of T2D and the frequent presence of additional CVD risk factors at the time of diagnosis, this implementation of PRS is unlikely to be of benefit in managing CVD risk in patients with T2D. They may, however, have a role predicting incident disease in patients with type 1 diabetes even when they are still adolescents or young adults; this will be important to test in future studies.

This study has several limitations. First, the data collected for this study come from an EHR-linked genomic and precision medicine cohort and does not include adjudicated disease status or outcomes which may have resulted in some degree of phenotype misclassification.

Second, the RECODe equation is designed to predict 10-year risk while the median follow-up in this study was 7 years. Since the main outcome of the study was a comparison of the RECODe with and without inclusion of PRS, these limitations would be expected to affect both models equally and therefore have negligible effects on their relative performance. Third, the indexing of entry into the analytic cohort to the time of diagnosis of T2D (rather than enrollment into PMBB) may have introduced survival bias as participants with T2D had to survive to enroll in PMBB and undergo genotyping; again, this would be expected to affect all models equally. Finally, the polygenic risk score in this study included low frequency and common variants (> 0.1%) and did not examine the predictive value of rare genetic variants known to affect CAD, HF and stroke risk.^31^ In conclusion, in the present study, we demonstrate that although CAD, HF and stroke PRS associate with CVD outcomes independent of traditional risk factors among individuals with T2D, the addition of PRS to contemporary clinical risk models does not specifically improve the predictive performance as compared to the baseline model.

## Data Availability

Raw data for the analysis dataset are not publicly available to preserve individuals privacy per the Health Insurance Portability and Accountability Act Privacy Rule.

## Data Availability

Raw data for the analysis dataset are not publicly available to preserve individuals’ privacy per the Health Insurance Portability and Accountability Act Privacy Rule.

## Acknowledgements

We acknowledge the Penn Medicine BioBank (PMBB) for providing data and thank the patient-participants of Penn Medicine who consented to participate in this research program. We would also like to thank the Penn Medicine BioBank team and Regeneron Genetics Center for providing genetic variant data for analysis. The PMBB is approved under IRB protocol# 813913 and supported by Perelman School of Medicine at University of Pennsylvania, a gift from the Smilow family, and the National Center for Advancing Translational Sciences of the National Institutes of Health under CTSA award number UL1TR001878.

## Author Contributions

Conceptualization: SMD, NLT. Methodology: SMD, NLT. Data generation and processing: SMD, NLT, RJ. Data visualization: NLT. Formal analysis: NLT. Project administration: NLT. Supervision: SMD, JC, BFV. Writing and crucial editing: SMD, NLT, RJ, MGL, GS, BFV, JC.

## Cited Sources

1. American Diabetes Association AD. 9. Cardiovascular Disease and Risk Management: Standards of Medical Care in Diabetes-2018. Diabetes Care. 2018;41(Suppl 1):S86–S104. doi:10.2337/dc18-S009

2. Smith NL, Maynard C. The burden of diabetes-associated cardiovascular hospitalizations in Veterans Administration (VA) and non-VA medical facilities. Diabetes Care. 2004;27 Suppl 2(suppl 2):B27–32. doi:10.2337/diacare.27.suppl_2.b27

3. Basu S, Sussman JB, Berkowitz SA, Hayward RA, Yudkin JS. Development and validation of Risk Equations for Complications Of type 2 Diabetes (RECODe) using individual participant data from randomised trials. Lancet Diabetes Endocrinol. 2017;5(10):788–798. doi:10.1016/S2213-8587(17)30221-8

4. Ripatti S, Tikkanen E, Orho-Melander M, et al. A multilocus genetic risk score for coronary heart disease: case-control and prospective cohort analyses. The Lancet. 2010;376(9750):1393–1400. doi:10.1016/S0140-6736(10)61267-6

5. Manikpurage HD, Eslami A, Perrot N, et al. Polygenic Risk Score for Coronary Artery Disease Improves the Prediction of Early-Onset Myocardial Infarction and Mortality in Men. Circ Genom Precis Med. 2021;14(6):697–708. doi:10.1161/CIRCGEN.121.003452

6. Khera A v., Chaffin M, Aragam KG, et al. Genome-wide polygenic scores for common diseases identify individuals with risk equivalent to monogenic mutations. Nature Genetics 2018 50:9. 2018;50(9):1219–1224. doi:10.1038/S41588-018-0183-Z

7. Thériault S, Lali R, Chong M, Velianou JL, Natarajan MK, Paré G. Polygenic Contribution in Individuals With Early-Onset Coronary Artery Disease. Circ Genom Precis Med. 2018;11(1):e001849. doi:10.1161/CIRCGEN.117.001849

8. Natarajan P, Young R, Stitziel NO, et al. Polygenic risk score identifies subgroup with higher burden of atherosclerosis and greater relative benefit from statin therapy in the primary prevention setting. Circulation. 2017;135(22):2091–2101. doi:10.1161/CIRCULATIONAHA.116.024436

9. Tada H, Melander O, Louie JZ, et al. Risk prediction by genetic risk scores for coronary heart disease is independent of self-reported family history. Eur Heart J. 2016;37(6):561–567. doi:10.1093/EURHEARTJ/EHV462

10. Khera A v., Emdin CA, Drake I, et al. Genetic Risk, Adherence to a Healthy Lifestyle, and Coronary Disease. New England Journal of Medicine. 2016;375(24):2349–2358. doi:10.1056/NEJMOA1605086/SUPPL_FILE/NEJMOA1605086_DISCLOSURES.PDF

11. Ding K, Bailey KR, Kullo IJ. Genotype-informed estimation of risk of coronary heart disease based on genome-wide association data linked to the electronic medical record. BMC Cardiovasc Disord. 2011;11(1):1–9. doi:10.1186/1471-2261-11-66/TABLES/4

12. Mega JL, Stitziel NO, Smith JG, et al. Genetic risk, coronary heart disease events, and the clinical benefit of statin therapy: an analysis of primary and secondary prevention trials. The Lancet. 2015;385(9984):2264–2271. doi:10.1016/S0140-6736(14)61730-X

13. Elliott J, Bodinier B, Bond TA, et al. Predictive Accuracy of a Polygenic Risk Score– Enhanced Prediction Model vs a Clinical Risk Score for Coronary Artery Disease. JAMA. 2020;323(7):636–645. doi:10.1001/JAMA.2019.22241

14. Mosley JD, Gupta DK, Tan J, et al. Predictive Accuracy of a Polygenic Risk Score Compared With a Clinical Risk Score for Incident Coronary Heart Disease. JAMA. 2020;323(7):627–635. doi:10.1001/JAMA.2019.21782

15. Kember RL, Merikangas AK, Verma SS, et al. Polygenic Risk of Psychiatric Disorders Exhibits Cross-trait Associations in Electronic Health Record Data From European Ancestry Individuals. Biol Psychiatry. Published online July 6, 2020. doi:10.1016/j.biopsych.2020.06.026

16. Levin MG, Tsao NL, Singhal P, et al. Genome-wide association and multi-trait analyses characterize the common genetic architecture of heart failure. Nature Communications 2022 13:1. 2022;13(1):1–15. doi:10.1038/s41467-022-34216-6

17. Tcheandjieu C, Zhu X, Hilliard AT, et al. Large-scale genome-wide association study of coronary artery disease in genetically diverse populations. Nature Medicine 2022 28:8. 2022;28(8):1679–1692. doi:10.1038/s41591-022-01891-3

18. Mishra A, Malik R, Hachiya T, et al. Stroke genetics informs drug discovery and risk prediction across ancestries. Nature 2022 611:7934. 2022;611(7934):115–123. doi:10.1038/s41586-022-05165-3

19. Rader DJ, Damrauer SM, Chaudhary K, et al. Association of the V122I Hereditary Transthyretin Amyloidosis Genetic Variant With Heart Failure Among Individuals of African or Hispanic/Latino Ancestry. JAMA. 2019;322(22):2191–2202. doi:10.1001/JAMA.2019.17935

20. Using Population Descriptors in Genetics and Genomics Research: A New Framework for an Evolving Field. Using Population Descriptors in Genetics and Genomics Research. Published online 2023. doi:10.17226/26902

21. Lyles CR, Harris LT, Jordan L, et al. Patient race/ethnicity and shared medical record use among diabetes patients. Med Care. 2012;50(5):434–440. doi:10.1097/MLR.0b013e318249d81

22. Klinger E V., Carlini S V., Gonzalez I, et al. Accuracy of Race, Ethnicity, and Language Preference in an Electronic Health Record. J Gen Intern Med. 2015;30(6):719. doi:10.1007/S11606-014-3102-8

23. Grambsch PM, Therneau TM. Proportional hazards tests and diagnostics based on weighted residuals. Biometrika. 1994;81(3):515–526. doi:10.1093/BIOMET/81.3.515

24. van Buuren S, Groothuis-Oudshoorn K. mice: Multivariate Imputation by Chained Equations in R. J Stat Softw. 2011;45(3):1–67. doi:10.18637/JSS.V045.I03

25. Liu D, Cai T, Zheng Y. Evaluating the Predictive Value of Biomarkers with Stratified Case-Cohort Design. Biometrics. 2012;68(4):1219. doi:10.1111/J.1541-0420.2012.01787.X

26. Pepe MS, Zheng Y, Jin Y, et al. Evaluating the ROC performance of markers for future events. Lifetime Data Analysis 2007 14:1. 2007;14(1):86–113. doi:10.1007/S10985-007-9073-X

27. 2019 ACC/AHA Guideline on the Primary Prevention of Cardiovascular Disease - American College of Cardiology. Accessed May 22, 2022. https://www.acc.org/latest-in-cardiology/ten-points-to-remember/2019/03/07/16/00/2019-acc-aha-guideline-on-primary-prevention-gl-prevention

28. Mosley JD, Gupta DK, Tan J, et al. Predictive Accuracy of a Polygenic Risk Score Compared With a Clinical Risk Score for Incident Coronary Heart Disease. JAMA. 2020;323(7):627–635. doi:10.1001/JAMA.2019.21782

29. Tzoulaki I, Liberopoulos G, Ioannidis JPA. Assessment of claims of improved prediction beyond the Framingham risk score. JAMA. 2009;302(21):2345–2352. doi:10.1001/JAMA.2009.1757

30. Ye Y, Chen X, Han J, Jiang W, Natarajan P, Zhao H. Interactions between Enhanced Polygenic Risk Scores and Lifestyle for Cardiovascular Disease, Diabetes, and Lipid Levels. Circ Genom Precis Med. 2021;14:3128. doi:10.1161/CIRCGEN.120.003128

31. Nikpay M, Goel A, Won HH, et al. A comprehensive 1000 Genomes–based genome-wide association meta-analysis of coronary artery disease. Nature Genetics 2015 47:10. 2015;47(10):1121–1130. doi:10.1038/ng.3396

