## Supplemental Information and Figures for "Evaluation of the Performance of the RECODe Equation with the Addition of Polygenic Risk Scores for Adverse Cardiovascular Outcomes in Individuals with Type II Diabetes"

**Penn Medicine BioBank Banner Author List and Contribution Statements**

**PMBB Leadership Team**

Daniel J. Rader, M.D., Marylyn D. Ritchie, Ph.D.

Contribution: All authors contributed to securing funding, study design and oversight. All authors reviewed the final version of the manuscript.

**Patient Recruitment and Regulatory Oversight**

JoEllen Weaver, Nawar Naseer, Ph.D., M.P.H., Afiya Poindexter, Khadijah Hu-Sain, Yi-An Ko, Ph.D.

Contributions: JW manages patient recruitment and regulatory oversight of study.  NN manages participant engagement, assists with regulatory oversight, and researcher access. AP, KH, YK perform recruitment and enrollment of study participants.

**Lab Operations**

JoEllen Weaver, Meghan Livingstone, Fred Vadivieso, Stephanie DerOhannessian, Teo Tran, Julia Stephanowski, Monica Zielinski, Ned Haubein, Joseph Dunn

Contribution: JW, ML, FV, SD conduct oversight of lab operations.  ML, FV, AK, SD, TT, JS, MZ perform sample processing. NH, JD are responsible for sample tracking and the laboratoryinformation management system.

**Clinical Informatics**

Anurag Verma, Ph.D., Colleen Morse Kripke, M.S. DPT, MSA, Marjorie Risman, M.S., Renae Judy, B.S.

Contribution: All authors contributed to the development and validation of clinical phenotypes used to identify study subjects and (when applicable) controls.

**Genome Informatics**

Anurag Verma Ph.D., Shefali S. Verma, Ph.D., Yuki Bradford, M.S., Scott Dudek, M.S., Theodore Drivas, M.D., Ph.D.

Contribution: A.V., S.S.V. are responsible for the analysis, design, and infrastructure needed to quality control genotype and exome data. Y.B. performs the analysis. T.D. and A.V. providesvariant and gene annotations and their functional interpretation of variants.

**Regeneron Genetics Center Banner Author List and Contribution Statements**

All authors are listed in alphabetical order.

**RGC Management and Leadership Team**

Goncalo Abecasis, Ph.D., Aris Baras, M.D., Michael Cantor, M.D., Giovanni Coppola, M.D., Aris Economides, Ph.D., John D. Overton, Ph.D., Jeffrey G. Reid, Ph.D., Alan Shuldiner, M.D.

Contribution: All authors contributed to securing funding, study design and oversight, and review and interpretation of data and results. All authors reviewed and contributed to the final version of the manuscript.

**Sequencing and Lab Operations**

Christina Beechert, Caitlin Forsythe, M.S., Erin D. Fuller, Zhenhua Gu, M.S., Michael Lattari, Alexander Lopez, M.S., John D. Overton, Ph.D., Thomas D. Schleicher, M.S., Maria Sotiropoulos Padilla, M.S., Karina Toledo, Louis Widom, Sarah E. Wolf, M.S., Manasi Pradhan, M.S., Kia Manoochehri, Ricardo H. Ulloa.

Contribution: C.B., C.F., K.T., A.L., and J.D.O. performed and are responsible for sample genotyping. C.B, C.F., E.D.F., M.L., M.S.P., K.T., L.W., S.E.W., A.L., and J.D.O. performed and are responsible for exome sequencing. T.D.S., Z.G., A.L., and J.D.O. conceived and are responsible for laboratory automation. M.P., K.M., R.U., and J.D.O are responsible for sample tracking and the library information management system.

**Genome Informatics**

Xiaodong Bai, Ph.D., Suganthi Balasubramanian, Ph.D., Leland Barnard, Ph.D., Andrew Blumenfeld, Yating Chai, Ph.D., Gisu Eom, Lukas Habegger, Ph.D., Young Hahn, Alicia Hawes, B.S., Shareef Khalid, Jeffrey G. Reid, Ph.D., Evan K. Maxwell, Ph.D., John Penn, M.S., Jeffrey C. Staples, Ph.D., Ashish Yadav, M.S.

Contribution: X.B., A.H., Y.C., J.P., and J.G.R. performed and are responsible for analysis needed to produce exome and genotype data. G.E., Y.H., and J.G.R. provided compute infrastructure development and operational support. S.K., S.B., and J.G.R. provide variant and gene annotations and their functional interpretation of variants. E.M., L.B., J.S., A.B., A.Y., L.H., J.G.R. conceived and are responsible for creating, developing, and deploying analysis platforms and computational methods for analyzing genomic data.

**Clinical Informatics**

Nilanjana Banerjee, Ph.D., Michael Cantor, M.D.

Contribution: All authors contributed to the development and validation of clinical phenotypes used to identify study subjects and (when applicable) controls.

**Analytical Genomics and Data Science**

Goncalo Abecasis, Ph.D., Amy Damask, Ph.D., Lauren Gurski, Alexander Li, Ph.D., Nan Lin, Ph.D., Daren Liu, Jonathan Marchini Ph.D., Anthony Marcketta, Shane McCarthy, Ph.D., Colm O’Dushlaine, Ph.D., Charles Paulding, Ph.D., Claudia Schurmann, Ph.D., Dylan Sun, Tanya Teslovich, Ph.D., Cristopher Van Hout, Ph.D., Bin Ye

Contribution: Development of statistical analysis plans. QC of genotype and phenotype files and generation of analysis ready datasets. Development of statistical genetics pipelines and tools and use thereof in generation of the association results. QC, review and interpretation of result. Generation and formatting of results for manuscript figures. Contributions to the final version of the manuscript.

**Therapeutic Area Genetics**

Jan Freudenberg, M.D., Nehal Gosalia, Ph.D., Claudia Gonzaga-Jauregui, Ph.D., Julie Horowitz, Ph.D., Kavita Praveen, Ph.D.

Contribution: Development of study design and analysis plans. Development and QC of phenotype definitions. QC, review, and interpretation of association results. Contributions to the final version of the manuscript.

**Planning, Strategy, and Operations**

Paloma M. Guzzardo, Ph.D., Marcus B. Jones, Ph.D., Lyndon J. Mitnaul, Ph.D.

Contribution: All authors contributed to the management and coordination of all research activities, planning and execution. All authors managed the review of data and results for the manuscript. All authors contributed to the review process for the final version of the manuscript.

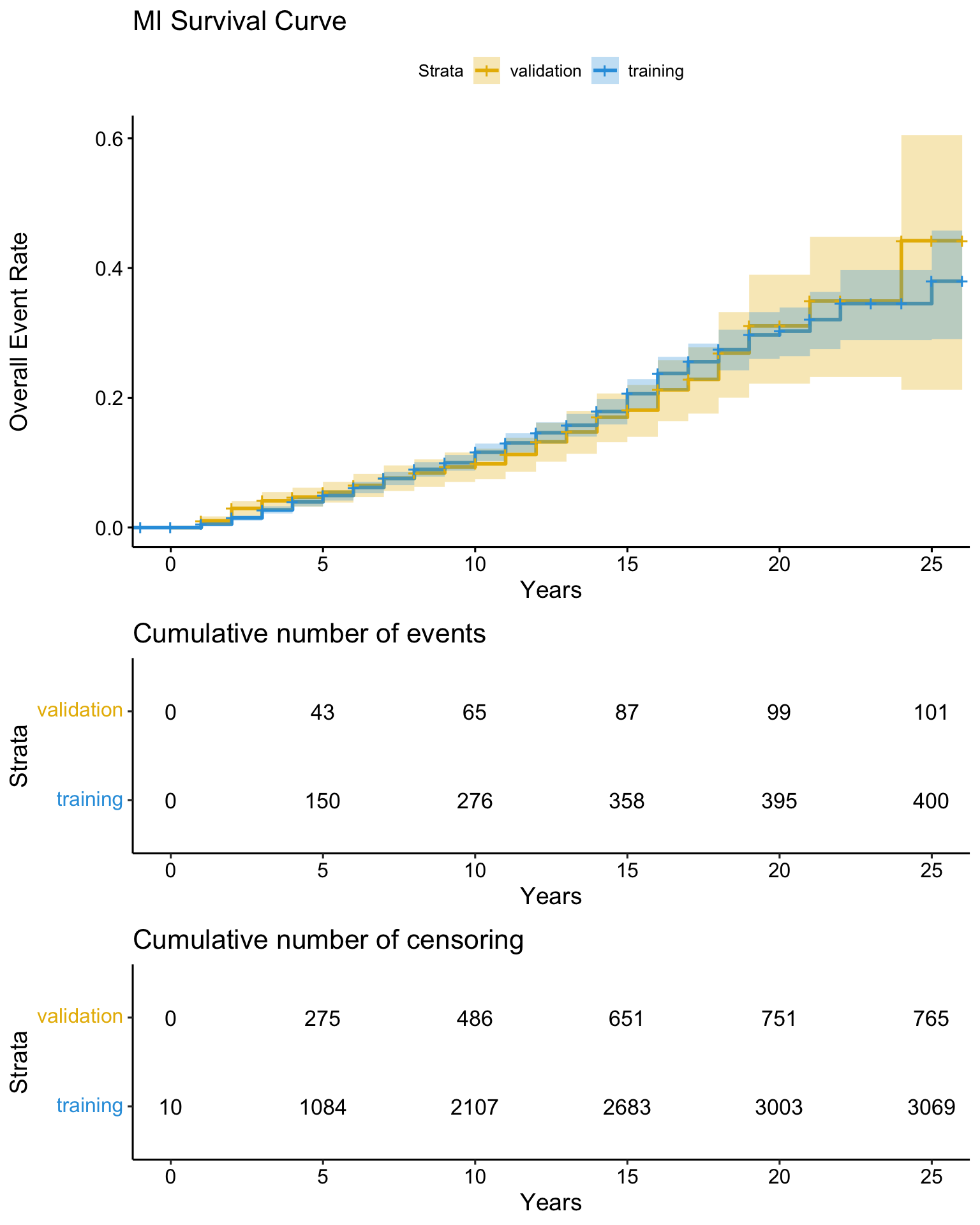

**Supplemental Figure 1) Unadjusted MI Event Rate Curve of event rate data to MI event (in months) after the diagnosis of T2D.** The number of events at 5, 10, 15, 20 and 25 years are shown in a table immediately below the survival curves.

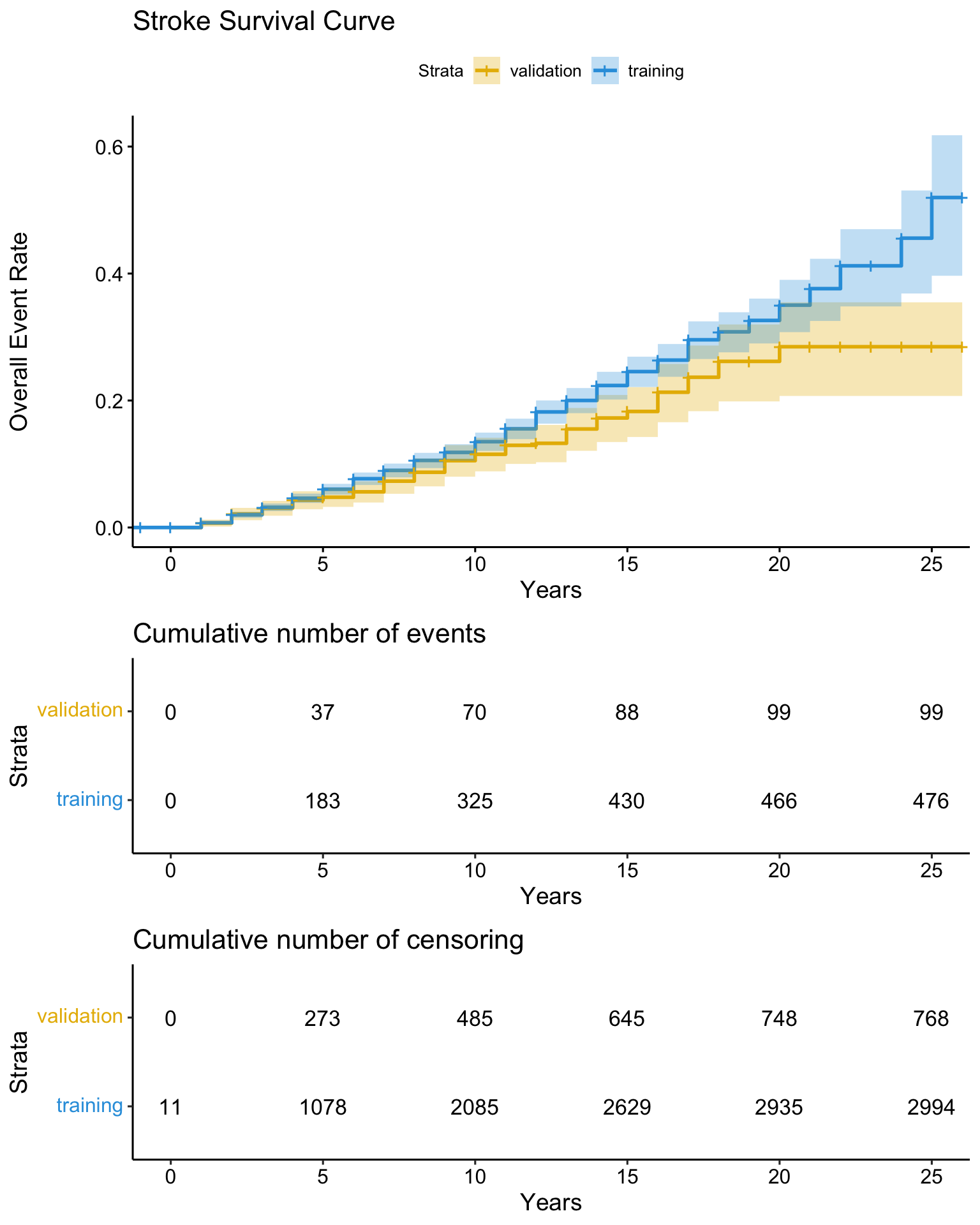

**Supplemental Figure 2) Unadjusted stroke event rate curve of event rate data to stroke event (in months) after the diagnosis of T2D.** The number of events at 5, 10, 15, 20 and 25 years are shown in a table immediately below the survival curves.

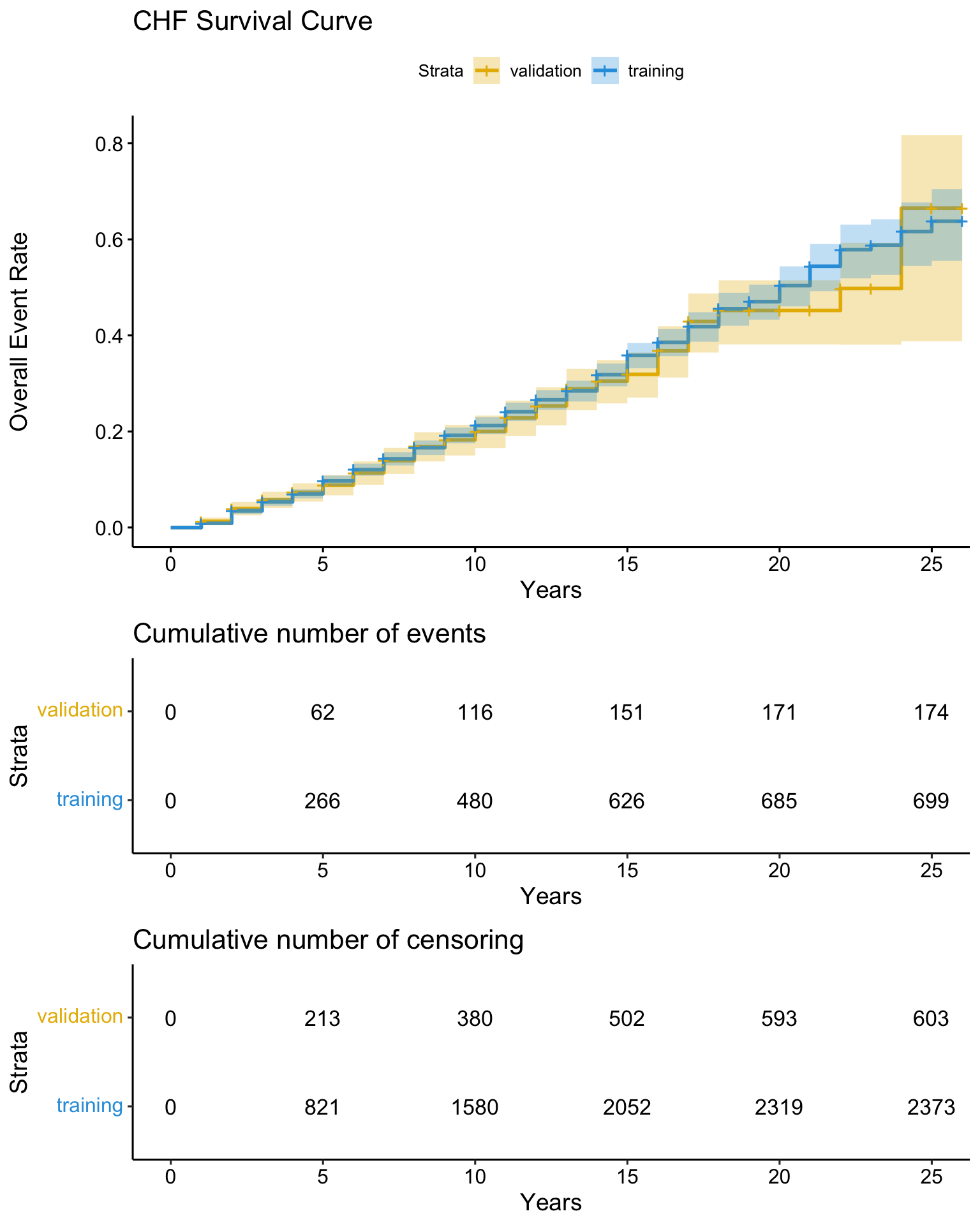

**Supplemental Figure 3) Unadjusted HF event rate rurve of event rate data to HF event (in months) after the diagnosis of T2D.** The number of events at 5, 10, 15, 20 and 25 years are shown in a table immediately below the survival curves.

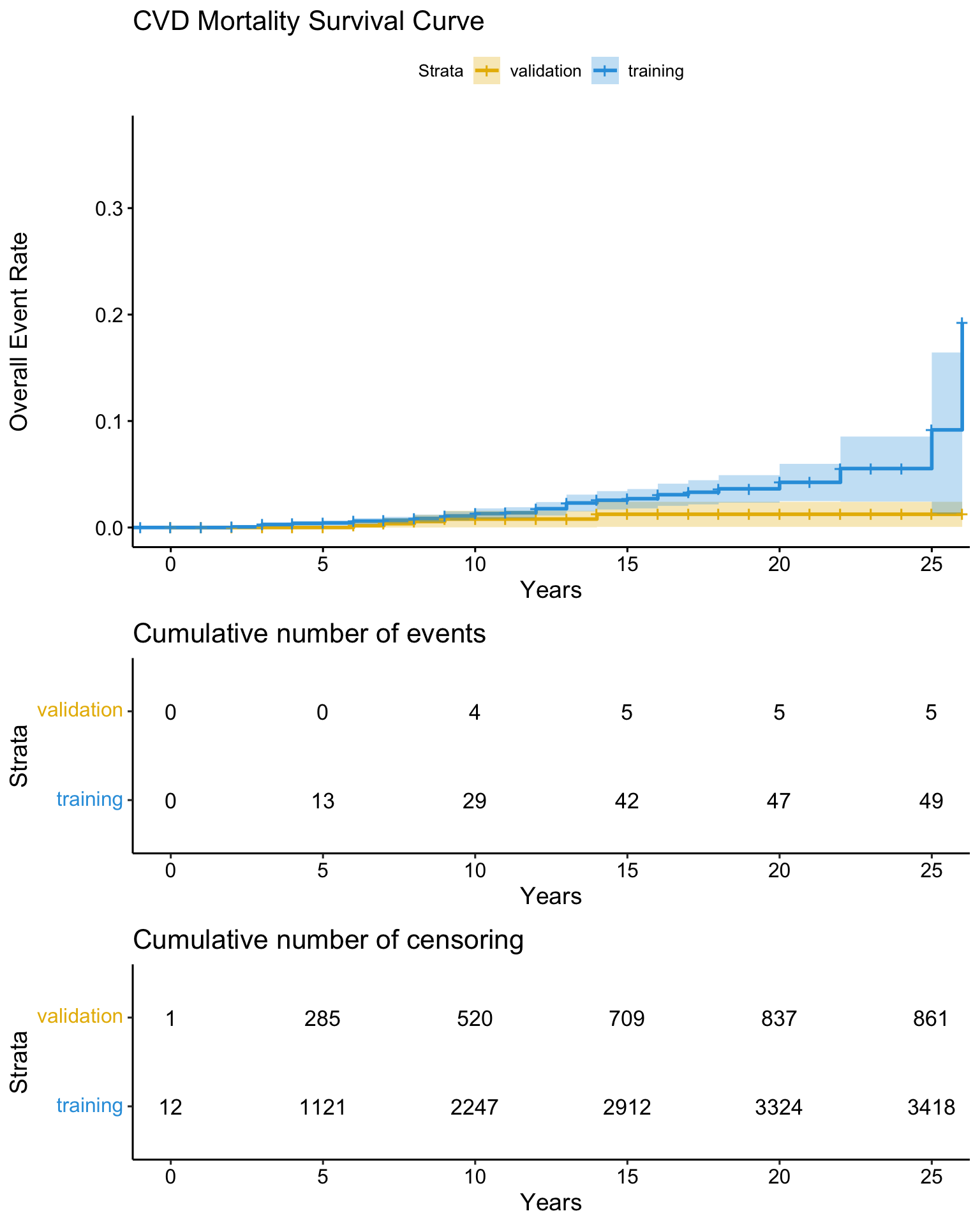

**Supplemental Figure 4) Unadjusted cardiovascular mortality event rate curve of event rate data to cardiovascular mortality event (in months) after the diagnosis of T2D.** The number of events at 5, 10, 15, 20 and 25 years are shown in a table immediately below the survival curves.

A)
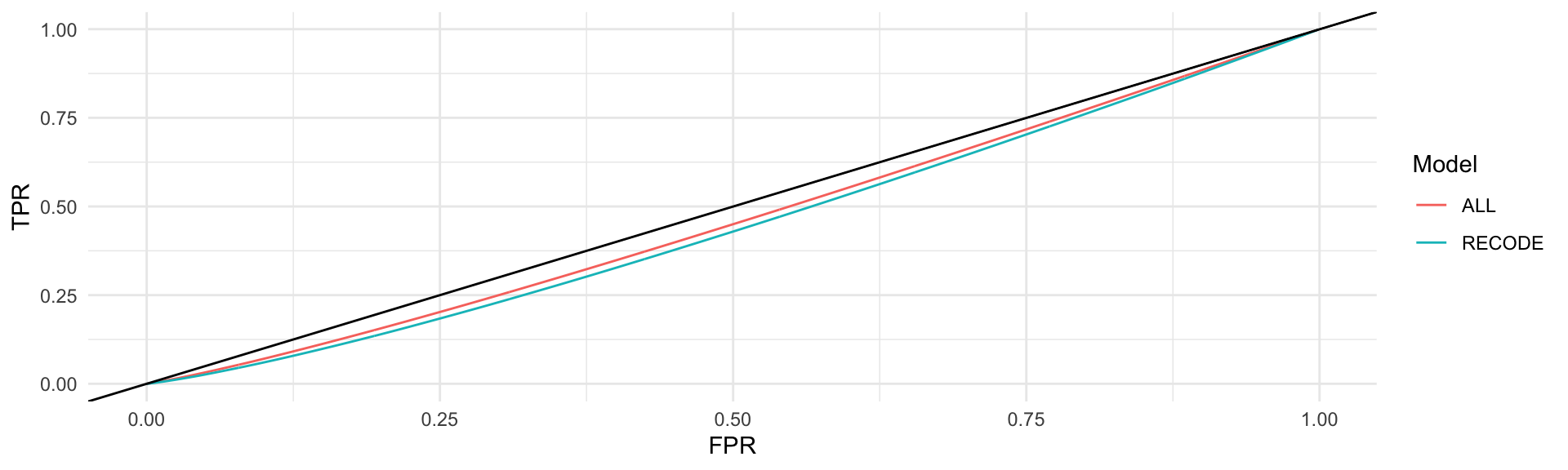

B)
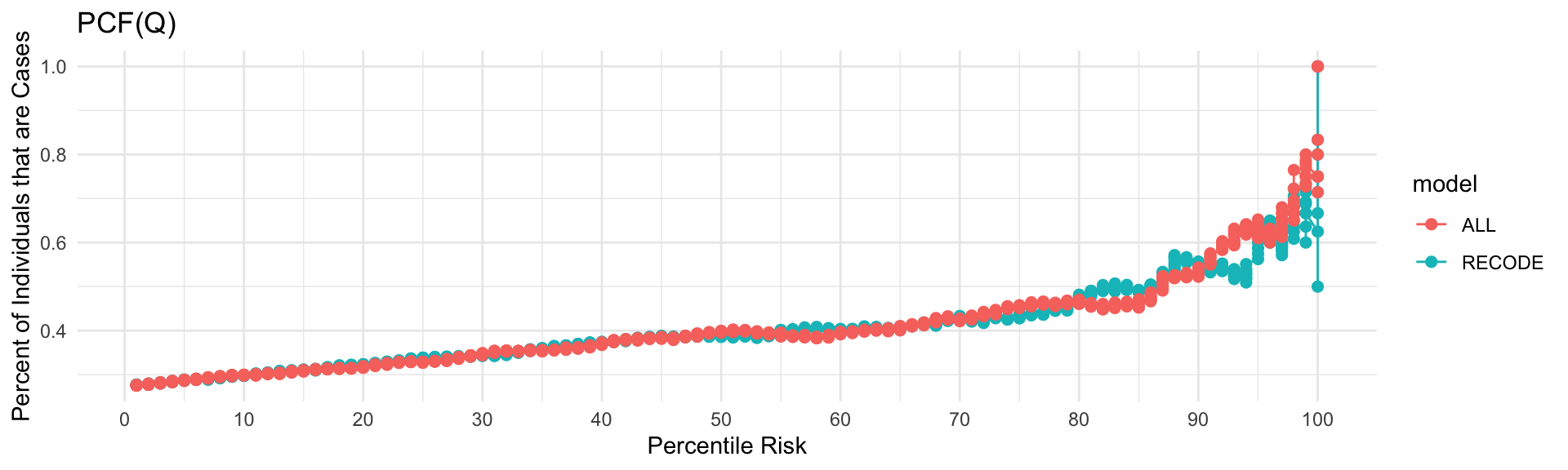

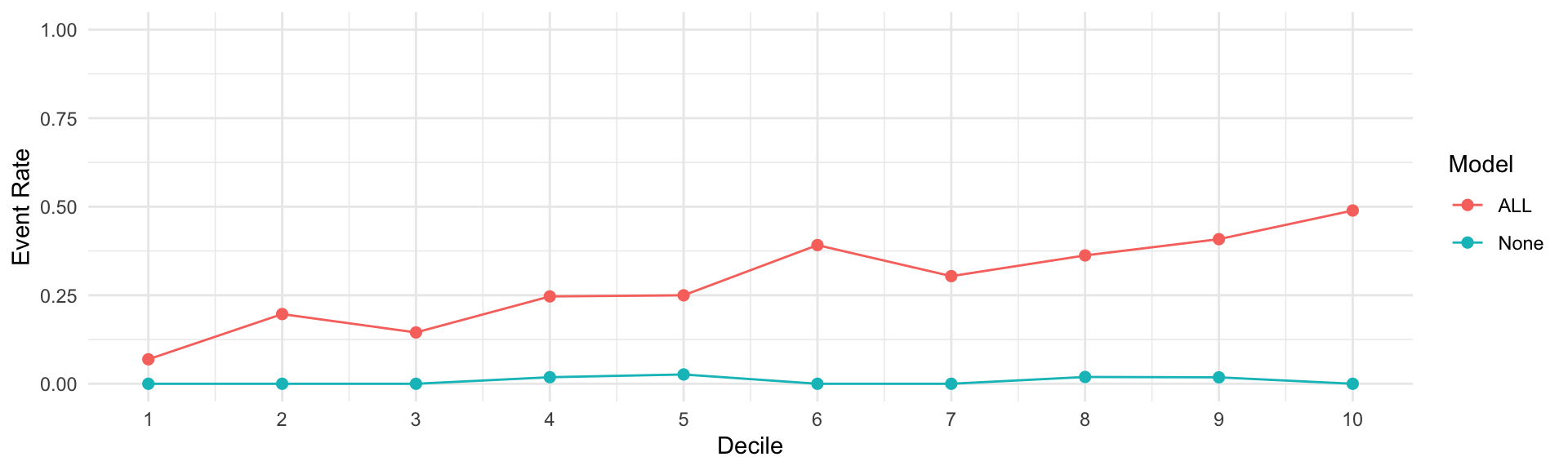

D)
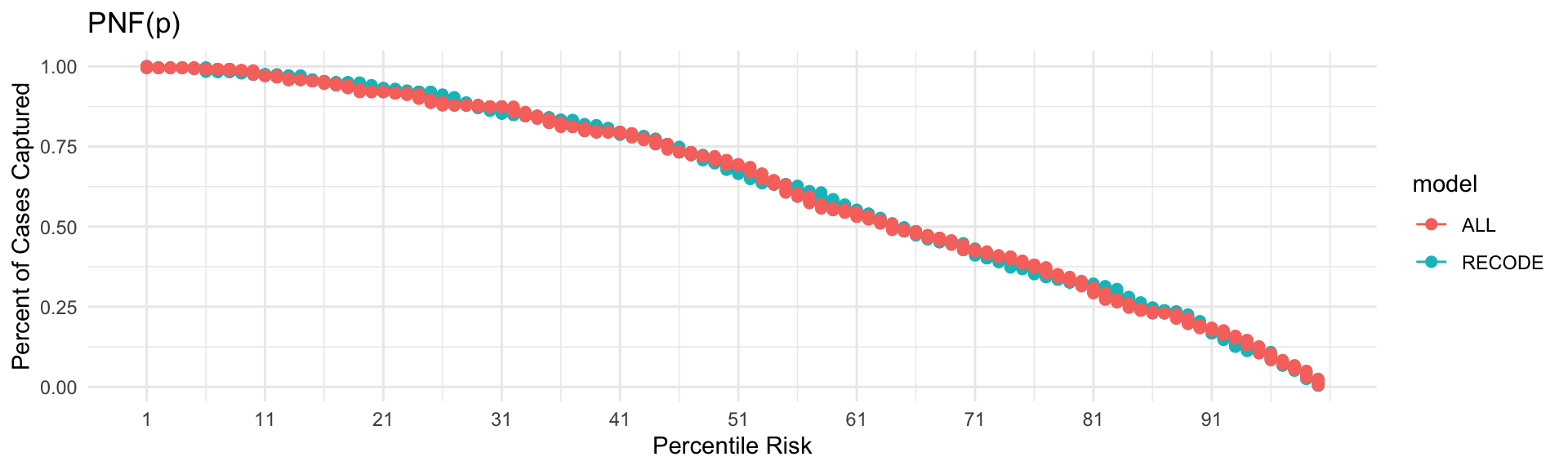

**Supplemental Figure 5) Performance of the RECODE Model for Cardiovascular Mortality with the addition of PRS** A) AUC, B) Event Rate, C) PCF(Q), D) PNF(P). The blue curve illustrates the corresponding curve modeled using the RECODe model without the addition of the PRS. The red curve illustrates the same curve modeled using the RECODe model with the addition of a CAD, Stroke and HF PRS.

A)
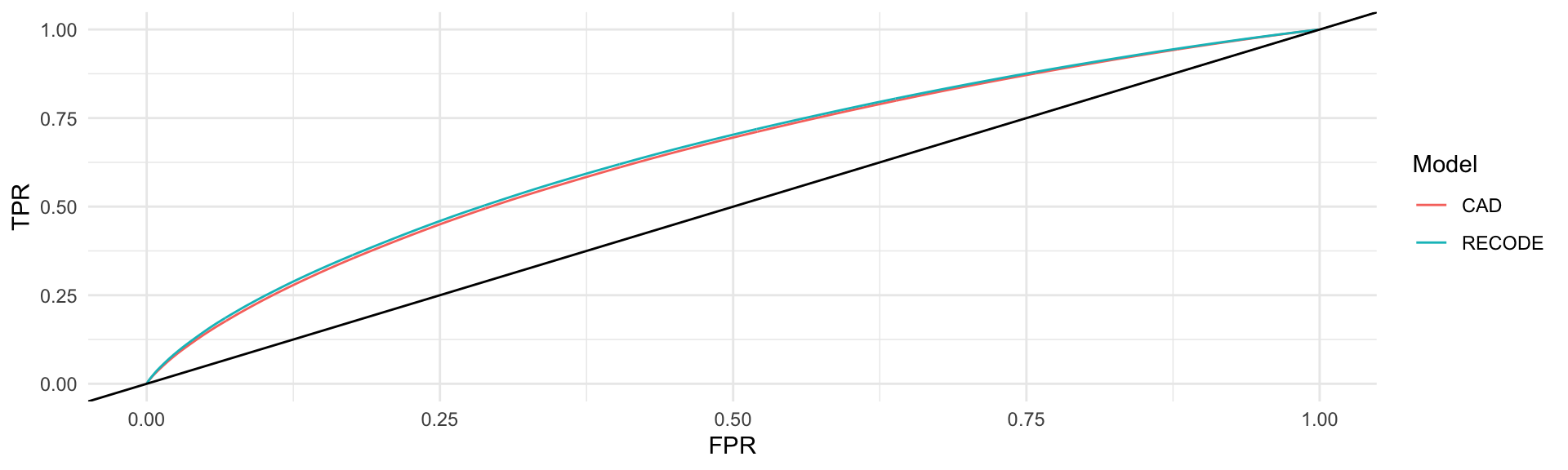

B)
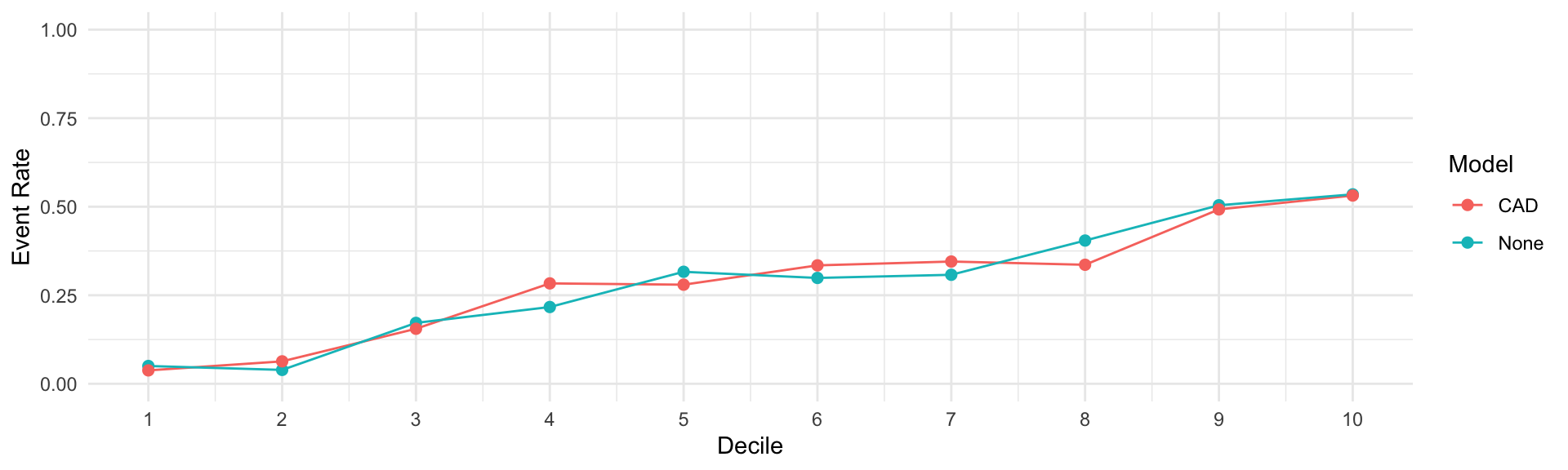

C)
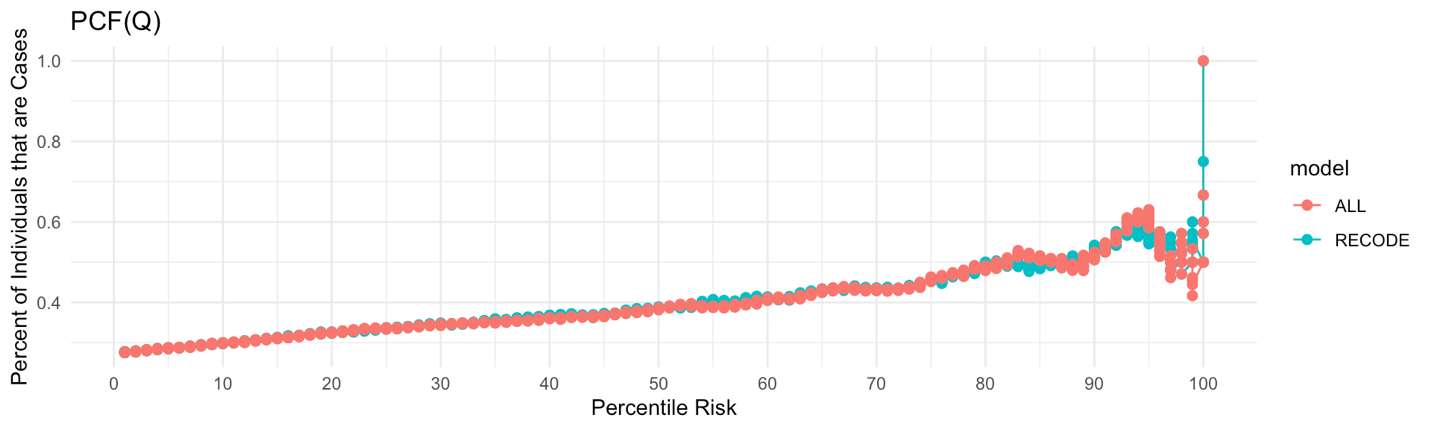

D)
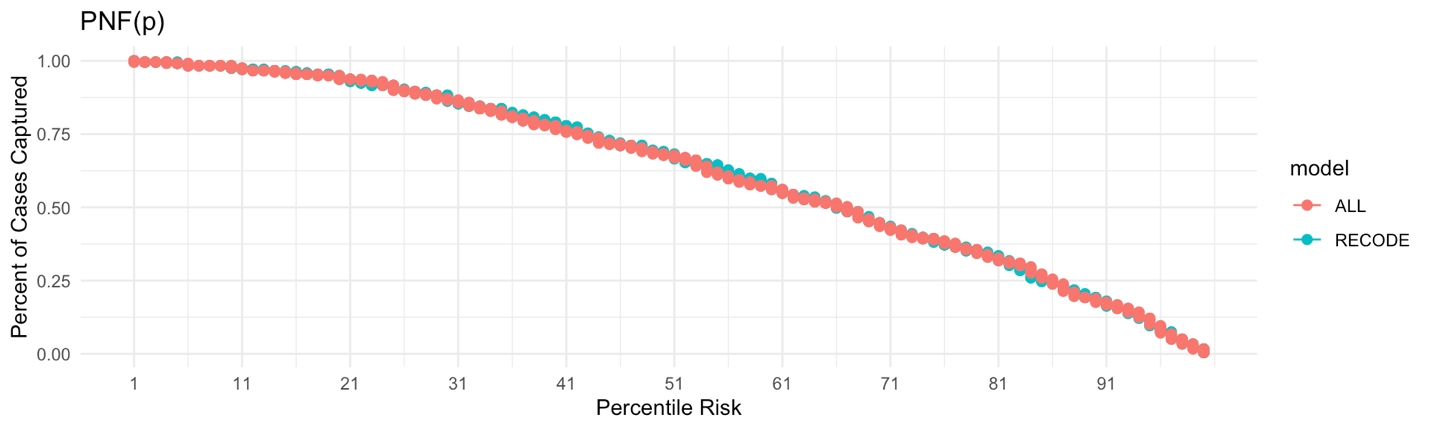

**Supplemental Figure 6) Performance of the RECODE Model for MI with the Addition of various PRS** A) AUC, B) Survival, C) PCF(Q), D) PNF(P). The blue curve illustrates the corresponding curve modeled using the RECODe model without the addition of the PRS. The red curve illustrates the same curve modeled using the RECODe model with the addition of a CAD, Stroke and HF PRS.

A)
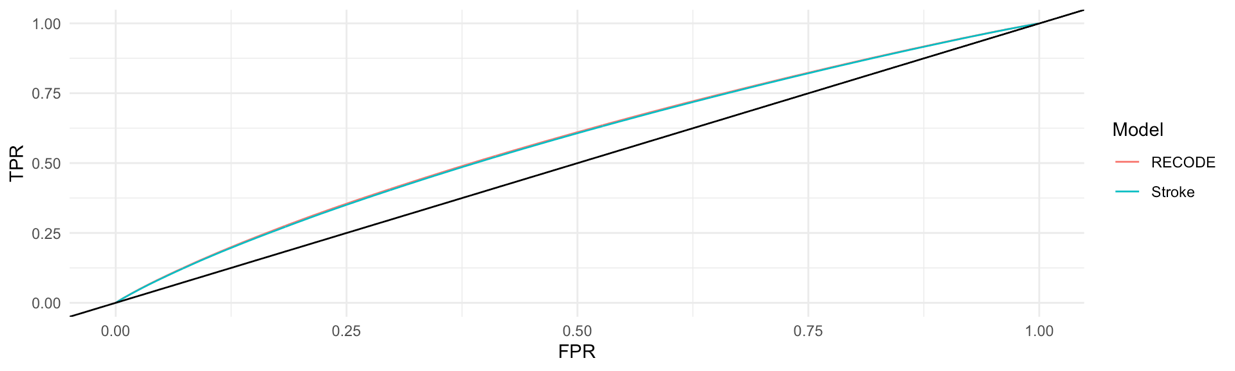

B)
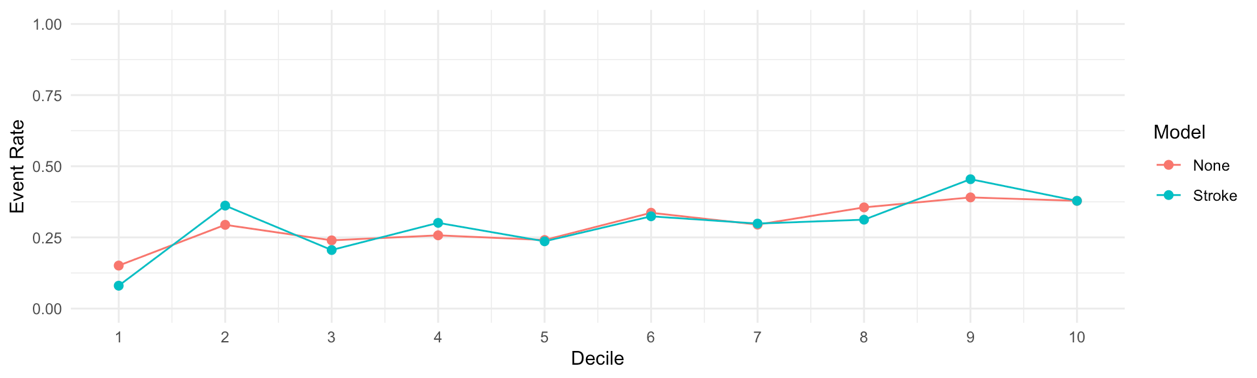

C)
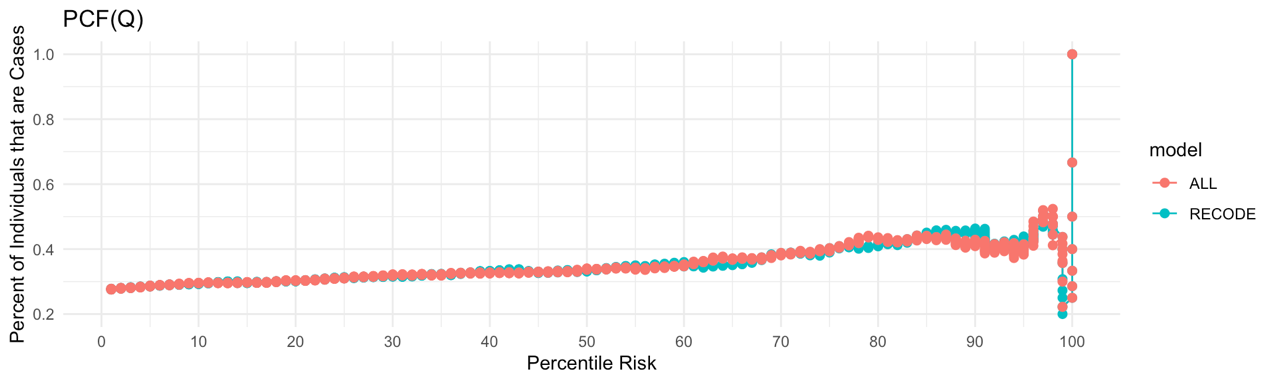

D)
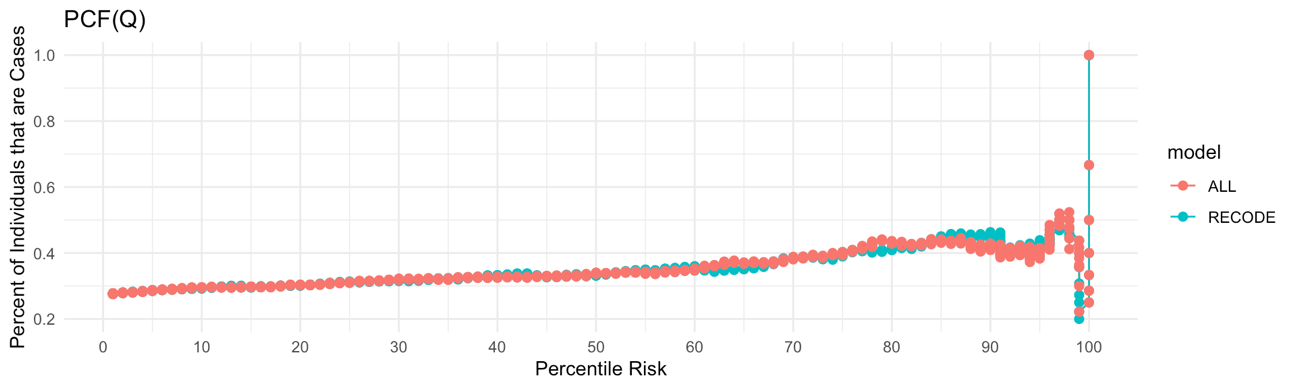

**Supplemental Figure 7) Performance of the RECODE Model for Stroke with the Addition of various PRS** A) AUC, B) Event Rate, C) PCF(Q), D) PNF(P). The blue curve illustrates the corresponding curve modeled using the RECODe model without the addition of the PRS. The red curve illustrates the same curve modeled using the RECODe model with the addition of a CAD, Stroke and HF PRS.

A)
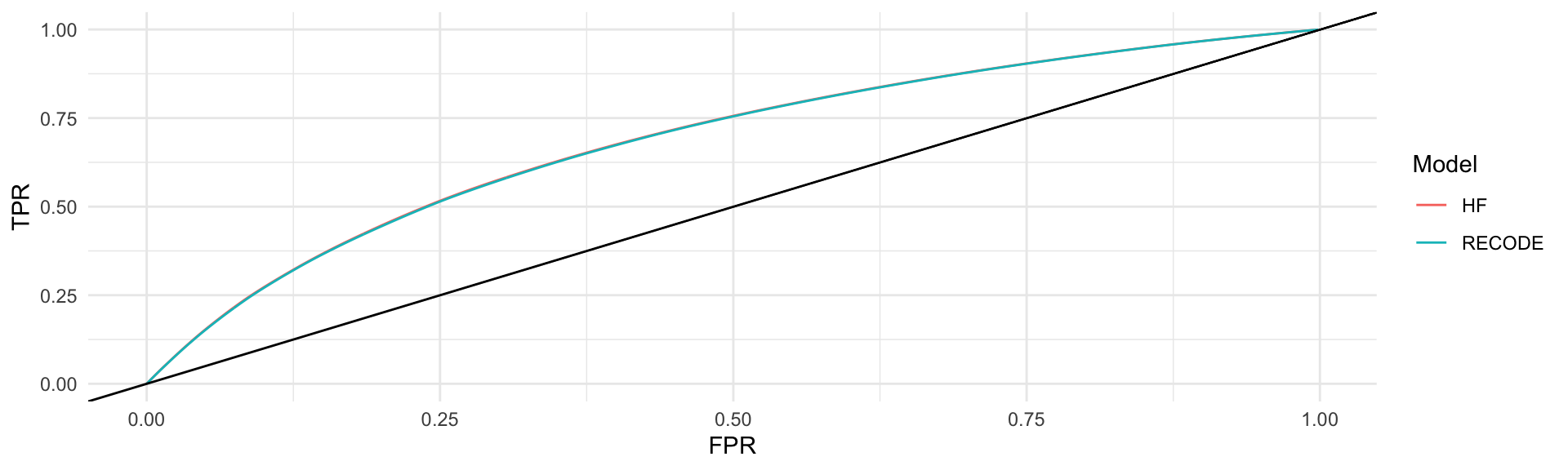

B)
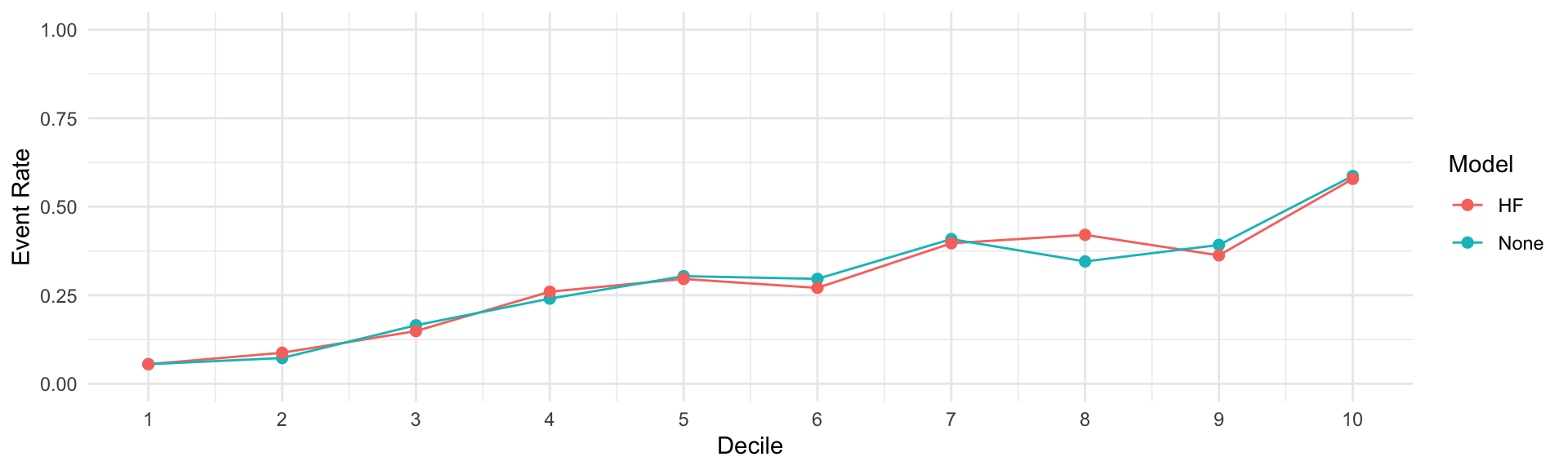

C)
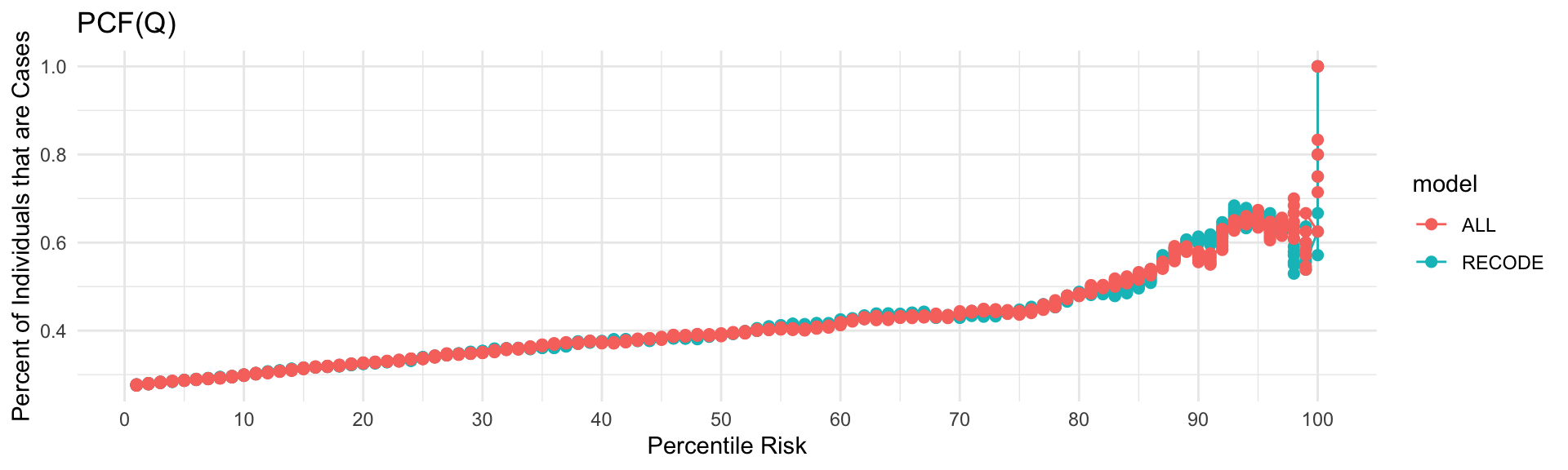

D)
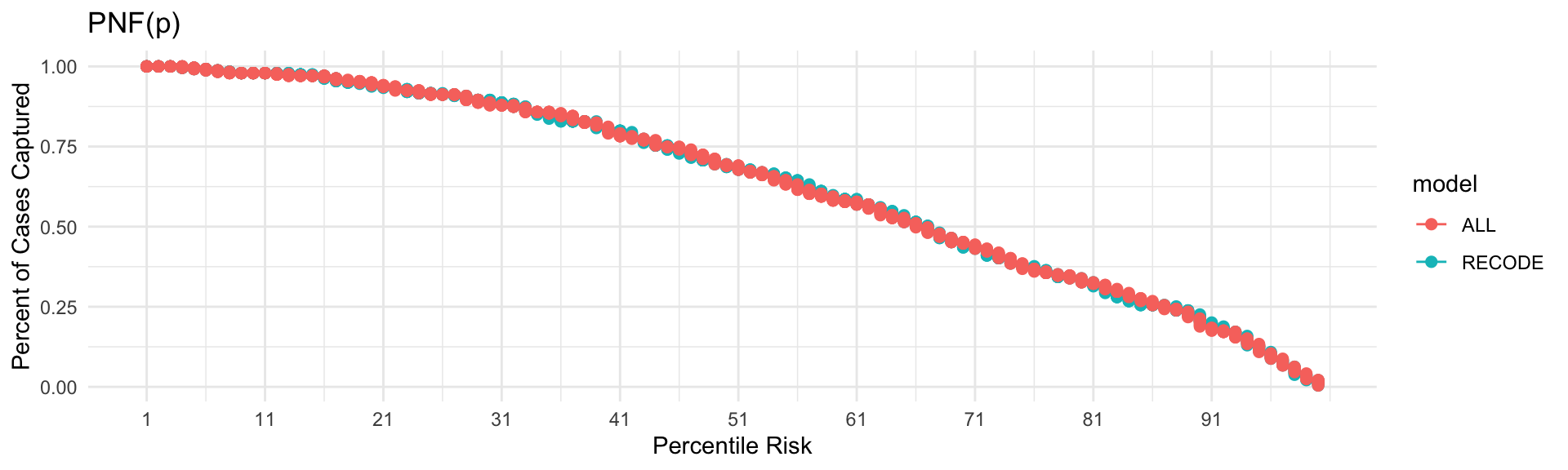

**Supplemental Figure 8) Performance of the RECODE Model for HF with the Addition of various PRS** A) AUC, B) Survival, C) PCF(Q), D) PNF(P). The blue curve illustrates the corresponding curve modeled using the RECODe model without the addition of the PRS. The red curve illustrates the same curve modeled using the RECODe model with the addition of a CAD, Stroke and HF PRS.
